# A multimodal AI model for modeling the genetic risk factor of Alzheimer’s disease

**DOI:** 10.64898/2026.04.13.26350803

**Authors:** Thong Nguyen, Carter Woods, Jian Liu, Chris Wang, Ai-Ling Lin, Jianlin Cheng

**Affiliations:** Department of Electrical Engineering and Computer Science, University of Missouri, Columbia, MO, USA; Roy Blunt NextGen Precision Health, University of Missouri, Columbia, MO, USA

**Author notes:** Corresponding author: Ai-Ling Lin,., Corresponding author: Jianlin Cheng,.

**Keywords:** Alzheimer’s disease, multimodal AI, transformer, contrastive learning, self-supervised learning, data integration

## Abstract

The apolipoprotein E *ε*4 (APOE4) allele is the strongest genetic risk factor for late-onset Alzheimer’s disease (AD), the most common form of dementia. APOE4 carriers exhibit cerebrovascular and metabolic dysfunction, structural brain alterations, and gut microbiome changes decades before the onset of clinical symptoms. Better understanding of the early manifestion of these physiological changes is critical for development of timely AD interventions and risk reduction protocols. Multi-modal datasets encompassing a wide range of APOE *ε*4 and AD associated biomarkers provide a valuable opportunity to gain insight into the APOE4 phenotype; however, these datasets often present analytical challenges due to small sample sizes and high heterogeneity. Here, we propose a two-stage multimodal AI model (APOEFormer) that integrates blood metabolites, brain vascular and structural MRI, microbiome profiles, and other clinical and demographic data to predict APOE4 allele status. In the first stage, modality-specific encoders are used to generate initial representa-tions of input data modalities, which are aligned in a shared latent space via self-supervised contrastive learning during pretraining. The contrastive learning objective encourages learning of informative and consistent representations across modalities through leveraging cross-modality relationships. In the second stage, the pretrained representations are used as inputs to a multimodal transformer that integrates information across modalities to predict a key AD-risk genetic variant (APOE4). Across 10 independent experimental runs with different train–validation–test splits, APOEFormer predicts whether an individual carries an APOE4 allele with an average prediction accuracy of 75%, demonstrating robust performance under limited sample sizes. Post hoc perturbation analysis of the predictive model revealed valuable insights into the driving components of the APOE4 phenotype— including key blood biomarkers and brain regions strongly associated with APOE4.

## I. Introduction

The apolipoprotein E *ε*4 (APOE4) allele is the strongest genetic risk factor associated with Alzheimer’s disease (AD). A single APOE4 allele increases an individual’s risk late-onset AD development by 3–7 times in heterozygous carriers while homogenous APOE4 carriers experience up to a 12-fold increase in risk of AD development [1], [2]. Beyond higher risk to AD development the APOE4 allele is associated with a complex cascade of physiological effects, including metabolic dysfunction, structural brain alterations, and gut microbiome changes that occur decades before and are believed to contribute to AD development. Understanding how these diverse biomarkers coalesce in the APOE4 phenotype is essential to identify early disease mechanisms and develop precision interventions.

Deconvolution of the complex APOE4 phenotype requires a sophisticated integration of multi-modal datasets, starting with the foundational role of magnetic resonance imaging (MRI) in capturing early neuroanatomical changes. In APOE4 carriers, MRI-derived measures frequently reveal accelerated cortical thinning and cerebral blood flow, both measures connected to neurodegeneration [3], [4]. Complementary non-imaging biomarkers—including biochemical measurements, microstructural indicators, and clinical variables—encode additional aspects of disease pathology that are not directly observable in imaging data [5], [6]. Moreover, recent research revealed that the bidirectional interaction between gut microbiome and brain plays an important role in AD [7].

Traditional machine learning approaches for multimodal phenotype analysis typically rely on handcrafted feature extraction followed by shallow fusion strategies, such as feature concatenation or ensemble classifiers [8]. While these methods have shown some success, they often struggle with high-dimensional inputs, limited sample sizes, and complex nonlinear interactions between modalities. Moreover, differences in scale, noise characteristics, and missing values across modalities can lead to poorly aligned representations and reduced generalization performance.

Deep learning methods have recently emerged as powerful alternatives for multimodal biomedical data integration. Convolutional neural networks have been widely applied to MRI-based AD classification [9], while multilayer perceptrons (MLPs) and autoencoder-based architectures have been used to model tabular clinical and biomarker data [10]. Despite these advances, many existing deep multimodal models are trained end-to-end using supervised objectives alone, making them vulnerable to overfitting in settings where labeled data is scarce or heterogeneous across cohorts.

Self-supervised and contrastive learning frameworks provide a principled approach to addressing these limitations by enabling representation learning from unlabeled or weakly labeled data. Contrastive objectives encourage embeddings of different views or modalities from the same subject to be similar, while maintaining discriminative separation across subjects [11], [12]. Such approaches have demonstrated strong performance in vision, language, and biomedical domains [13], particularly when annotations are limited or expensive to obtain. In multimodal medical settings, contrastive learning has been shown to effectively align imaging and non-imaging representations into a shared latent space, improving downstream predictive performance and robustness [14].

In parallel, transformer-based architectures have become increasingly influential due to their ability to model complex dependencies using attention mechanisms [15]. Transformers allow models to dynamically weight informative features and capture long-range interactions without relying on fixed receptive fields [16]. These properties are particularly advantageous for biomedical data, where disease-relevant signals may be distributed across spatially distant brain regions or across heterogeneous biomarker sets. Recent studies have successfully applied transformer-based models to medical imaging, clinical time series, and multimodal health data, demonstrating improved flexibility and interpretability compared to conventional architectures [17], [18].

In this work, we introduce a two-stage multimodal AI framework (APOEFormer) for APOE4 carrier prediction and phenotype characterization that combines contrastive representation learning with attention-based multimodal integration. In the first stage, modality-specific encoders are used to generate the representations of data modalities, which are aligned in a unified latent space by contrastive learning during self-supervised pretraining. This stage enables the model to learn robust, aligned representations of the heterogeneous modalities that capture phenotypical features without relying on labeled data. In the second stage, the pretrained representations are used as inputs to a transformer-based model that integrates information across modalities to predict whether a subject carries at least one APOE4 allele. Because the input representations are of high quality, fine-tuning the transformer model only needs a small number of labeled data samples.

By decoupling the representation learning from downstream predictive modeling, the proposed framework addresses key challenges in prediction of genetic status from multimodal biomedical data, including data heterogeneity, label scarcity, and complex cross-modal interactions. Moreover, the prediction of the AI model is further interpreted by the post hoc perturbation technique to identify key modalities and features associated with APOE4 variant, providing valuable insights about AD vulnerability. Therefore, APOEFormer provides a scalable and extensible approach to multimodal learning for genotype prediction and characterization that can generalize to a broader class of disease-associated biomarker classification tasks.

## II. Methods

### A. APOEFormer overview

APOEFormer is a multimodal AI framework designed to integrate heterogeneous subject data, such including neuroimaging and clinical biomarkers, into a unified representation for predicting an individual’s APOE4 carrier status.

The architecture of APOEFormer (Figure 1) follows a two-stage learning paradigm that decouples representation learning from downstream prediction tasks. In the first stage, modality-specific encoders are trained using a contrastive learning objective to project imaging and non-imaging data modalities into a shared latent space. Structural MRI volumes and perfusion images are encoded using a CLIP-based vision encoder [13], leveraging pretrained visual representations that have demonstrated strong transferability to medical imaging tasks [14], [19]. In parallel, heterogeneous non-imaging modalities—including microstructural features, multiple biomarker panels, and numeric clinical variables—are encoded using multilayer perceptron (MLP)–based networks. This design allows each modality to preserve domain-specific inductive biases while producing metrically aligned embeddings through self-superised contrastive learning [11], [12].

**Fig. 1.**
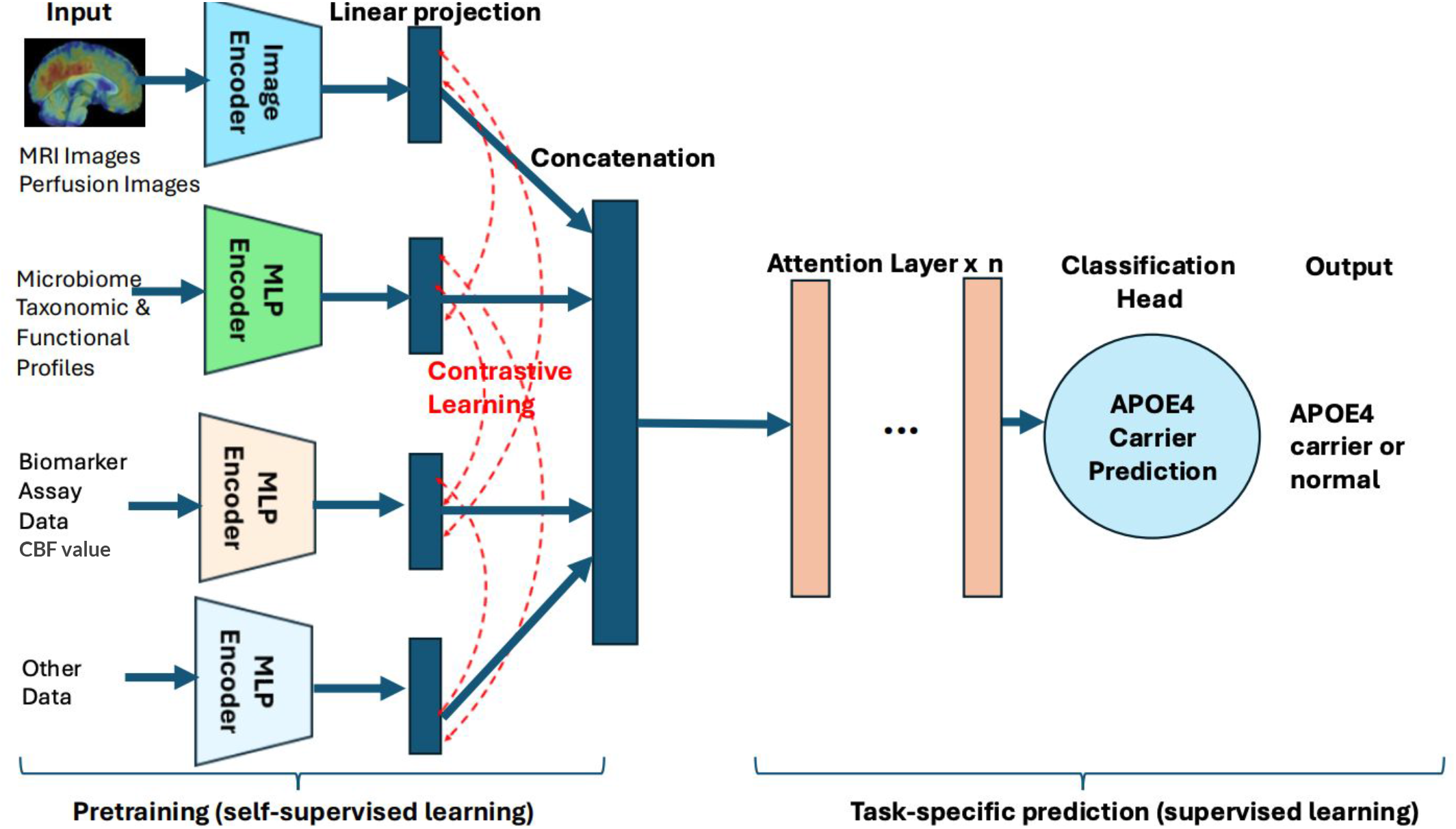
Overview of APOEFormer - a two-stage multimodal AI framework for Alzheimer’s disease analysis. Modality-specific encoders are first aligned using contrastive learning to produce shared latent representations from imaging and non-imaging data, which are then integrated by a transformer-based fusion module to predict the presence of an APOE4 allele(APOE4 carrier or not).

The self-supervised contrastive pretraining is particularly well suited to phenotype modeling because it does not require labels, where labeled outcomes are often limited, heterogeneous across cohorts, or affected by diagnostic uncertainty [20]. By aligning multimodal embeddings within the same subject and across subjects, the model learns improved representations of heterogeneous data modalities by leveraging modality relationships, while remaining robust to noise and missing data. The resulting embeddings serve as standardized representations for downstream modeling, analogous to representation alignment strategies used in self-supervised language and vision transformers [13], [21].

In the second stage, APOEFormer employs a transformer-based fusion module to integrate pretrained embeddings across modalities. Attention mechanisms enable the model to dynamically capture interactions among imaging and non-imaging features, allowing it to adaptively weight informative signals across modalities [16]. This flexibility is especially important in phenotype modeling, where the relative contributions of structural brain changes and non-imaging biomarkers may vary across individuals and disease stages.

The transformer encoder produces contextualized subject-level representations that summarize multimodal disease information. These representations are aggregated through attention-based pooling and passed to a prediction head that outputs a binary classification indicating whether a given patient carries the APOE4 allele. APOE4 is the strongest known genetic risk factor for late-onset AD and is associated with accelerated neurodegeneration, altered biomarker profiles, and increased disease susceptibility [22]. Predicting APOE4 carrier status from multimodal phenotypic data provides a biologically grounded and clinically meaningful target that links genetic risk to observable neuroimaging and clinical patterns.

By integrating contrastive self-supervised learning with transformer-based multimodal modeling, APOEFormer addresses key challenges in phenotype analysis, including data heterogeneity, limited labels, and complex cross-modal interactions. The proposed framework provides a scalable and extensible foundation for studying genetic risk variants and their effects in AD and can be generalized to study other diseases with multimodal data.

The components of APOEFormer, learning strategies, and methods to interpret predictions are described in the subsections below.

### B. Image encoder for encoding MRI images

Structural MRI and Perfusion image volumes provide rich anatomical information but pose challenges for representation learning due to their high dimensionality and variability in slice count across subjects. Direct 3D convolutional modeling is often data-intensive and sensitive to limited cohort sizes [23]. To address these challenges, we adopt a slice-wise encoding strategy that leverages large-scale pretrained visual representations generated by CLIP ViT-B/32 [19] while preserving depth-wise anatomical structure. Each MRI and Perfusion volume is decomposed along the axial dimension into an ordered sequence of 2D slices, enabling the use of powerful 2D vision transformers without discarding volumetric context.

Each slice undergoes intensity normalization to mitigate scanner-dependent variability and numerical instability arising from low-contrast regions or missing values. The normalized slices are converted into three-channel images and optionally augmented during training to improve robustness. Feature extraction is performed using the vision encoder of CLIP ViT-B/32, initialized from large-scale image–text pretraining [19]. Given the limited size typical of biomedical datasets, most CLIP parameters are frozen to stabilize optimization, while only the final transformer blocks are unfrozen to allow controlled task-specific adaptation. A gradual unfreezing strategy is employed, enabling additional layers to be released when validation performance plateaus, thereby balancing representation flexibility and generalization.

The high-dimensional CLIP embeddings are then projected into a compact latent space using a lightweight projection head composed of linear transformations, normalization, and nonlinear activation. Both the CLIP features and projected embeddings are 𝓁_2_-normalized to enforce consistent scale across slices and modalities. The slice encoding process can be summarized as

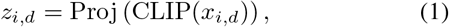

where *x*_*i,d*_ denotes the *d*-th slice of subject *i* and *z*_*i,d*_ is the corresponding slice embedding. The resulting embeddings form a depth-ordered sequence that is padded or truncated to enable batch-wise processing while preserving relative slice structure.

### C. MLP encoder for non-image modalities

In addition to imaging data, our framework incorporates multiple non-imaging modalities, including gut microbiome profiles, CBF value, blood-based biomarker panels, auxiliary clinical variables (e.g., gender, age, and body mass index [BMI]). These modalities encode complementary biological and phenotypic information but differ substantially in dimensionality, statistical distribution, and semantic meaning. Direct concatenation of such heterogeneous inputs can lead to scale imbalance and suboptimal feature interactions, particularly in attention-based fusion architectures [8].

To address this challenge, each non-imaging modality is processed using a dedicated multilayer perceptron (MLP) encoder that maps raw inputs into a latent embedding space. Each encoder consists of stacked transformation blocks comprising a linear projection, layer normalization, nonlinear activation, and dropout. Layer normalization is used to reduce covariate shift and improve training stability across modalities, while dropout provides regularization against overfitting [24]. Residual connections are applied whenever dimensionality permits, allowing the encoder to preserve informative low-level signals while learning higher-level abstractions.

A final normalization step followed by 𝓁_2_-normalization ensures that all modality embeddings lie on a comparable scale, facilitating stable cross-modal interaction during attention-based fusion. For a given modality input vector *x*_*i*_, the encoding process can be written as

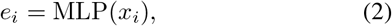

where *e*_*i*_ denotes the normalized modality embedding. By enforcing a consistent embedding dimensionality and normalization scheme across modalities, these MLP encoders provide a principled interface for integrating heterogeneous biomedical data.

### D. Contrastive Learning for Modality Alignment during Pretraining

Learning robust representations from heterogeneous biomedical data is challenging due to substantial variation in scale, noise characteristics, and semantic meaning across modalities. In multimodal clinical settings, imaging-derived features, molecular profiles, and structured clinical variables provide complementary but weakly aligned views of the same subject. To encourage coherent cross-modal representations prior to transformer fusion, we employ a contrastive learning-based pretraining strategy that explicitly aligns modality-specific embeddings at the subject level (Figure 1). During pretraining, each subject is represented by multiple modality embeddings, including imaging-based embeddings and non-imaging embeddings produced by modality-specific MLP encoders. All embeddings are normalized to lie in a shared metric space, enabling direct similarity comparisons across modalities. The contrastive objective follows an InfoNCE-style formulation, in which subject identity provides supervision: embeddings derived from different modalities of the same subject are treated as positive pairs, while embeddings originating from different subjects are treated as negatives [11], regardless of modality. This results in a multi-positive contrastive setting, where each anchor embedding is contrasted against all other modality embeddings within the minibatch.

Pairwise similarities are computed using temperature-scaled cosine similarity, and the InfoNCE normalization encourages embeddings associated with the same subject to form compact clusters while maintaining separation between subjects in the latent space. By aligning heterogeneous modalities through this objective, the model learns modality-invariant representations that capture subject-specific characteristics rather than modality-specific artifacts.

Formally, the contrastive objective is defined as

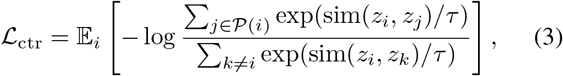

where *z*_*i*_ denotes a modality-specific embedding, 𝒫 (*i*) is the set of embeddings corresponding to the same subject (i.e., positive pairs), sim(*·,·*) denotes cosine similarity, and *τ* is a temperature hyperparameter controlling the sharpness of the similarity distribution. The expectation 𝔼_*i*_ denotes averaging over all anchor embeddings *i* within a mini-batch.

The objective ℒ_ctr_ is minimized during training. Minimizing this loss increases similarity between embeddings belonging to the same subject (maximizing the numerator) while reducing similarity to embeddings from different subjects (implicitly minimizing the denominator’s relative contribution), thereby promoting intra-subject compactness and inter-subject separation in the latent space.

This contrastive pretraining phase produces a shared embedding space in which representations from different modalities are aligned at the subject level, providing a robust initialization for subsequent transformer-based multimodal fusion and downstream prediction.

### E. Multimodal Transformer for Predicting Genotypes

To integrate the aligned representations of multiple modalities into a unified subject-level representation to predict APOE4 carrier status, we employ a transformer built on stacked self-attention layers [16] (Figure 1). All modality embeddings, including MRI and Perfusion images slice embeddings and static tabular embeddings, are treated as tokens within a single sequence. This formulation allows the model to learn context-dependent interactions across slices and modalities, capturing dependencies that are difficult to model using fixed aggregation strategies.

Each self-attention layer applies multi-head attention followed by dropout, residual connections, and layer normalization. This architecture enables stable gradient flow and progressively refines token representations by allowing each embedding to attend to all others. By stacking multiple attention layers, the transformer can capture higher-order cross-modal interactions and dynamically reweight modalities based on their relevance to the prediction task.

Following attention-based contextualization, the variable-length token sequence is compressed into a fixed-dimensional subject representation using a learned attention pooling mechanism [25]. Each token is assigned an importance score, and the final representation is computed as a weighted sum of token embeddings:

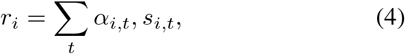

where *s*_*i,t*_ denotes the contextualized embedding of token *t* for subject *i*, and *α*_*i,t*_ are learned attention weights. The pooled representation is subsequently passed through a lightweight MLP to produce the final prediction. This combination of self-attention and learned pooling enables the model to emphasize relevant slices or modalities while suppressing less informative inputs.

### F. Supervised Transformer Training

Following the contrastive multimodal pretraining, the model is trained in a supervised manner to predict APOE4 carrier status at the subject level. In this phase, the pretrained modality encoders generate fixed-dimensional embeddings for each subject, which are then provided as input tokens to the transformer classifier. The transformer serves as the primary decision module, learning to integrate information across modalities and to produce a discriminative subject-level representation tailored to APOE *ε*4 prediction.

Supervised training is conducted using labeled subjects, with strict separation of training, validation, and test sets to prevent information leakage across individuals. For each subject, multimodal embeddings are assembled into a token sequence and processed by a stack of self-attention layers. Residual connections, dropout, and layer normalization are applied throughout the transformer, enabling stable optimization and allowing the model to dynamically reweight modalities based on their relevance to APOE4 status.

To address class imbalance and heterogeneous prediction difficulty inherent to APOE4 classification, the transformer is optimized using a focal loss objective [26]. Focal loss extends binary cross-entropy (BCE) by introducing multiplicative weighting factors that reduce the contribution of well-classified samples while emphasizing harder examples. This property is particularly important in clinical genetics settings where carrier prevalence is uneven.

Specifically, the supervised training objective is defined as

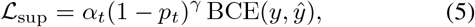

where *y ∈ {*0, 1*}* denotes APOE4 carrier status, *ŷ* is the predicted probability, and the binary cross-entropy term is given by

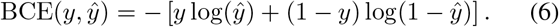

Here, the three components in Equation 5 are multiplicative rather than additive: BCE(*y, ŷ*) provides the base binary classification loss, *α*_*t*_ is a class-balancing coefficient that adjusts the relative weight of carriers and non-carriers, and the factor (1 − *ŷ*)^*γ*^ modulates the loss such that confidently classified samples contribute less to the overall objective. The focusing parameter *γ* controls the strength of this modulation.

Training is performed using the AdamW optimizer with weight decay, gradient clipping, and mixed-precision arithmetic to ensure numerical stability and computational efficiency. Early stopping based on validation loss is employed to mitigate overfitting, and the best-performing transformer model is selected for final evaluation.

### G. Multimodal Input Data

The proposed framework integrates multimodal input data spanning neuroimaging, molecular, microbial, and clinical domains to comprehensively characterize systemic and brain-specific biological processes. Structural MRI features were derived from high-resolution T1-weighted images, which provide detailed anatomical characterization of cortical thickness, subcortical volumes, and gray–white matter boundaries. Structural MRI has been widely used for quantifying neurodegeneration and regional atrophy patterns in aging and neurological disorders [27]. In addition to structural anatomy, functional perfusion information was incorporated through perfusion image, obtained using arterial spin labeling (ASL). ASL is a non-invasive MRI technique that quantifies cerebral blood flow (CBF) by magnetically labeling inflowing arterial blood water, enabling voxel-wise perfusion mapping without contrast agents [28].

To clearly distinguish image-based perfusion representations from derived quantitative summaries, we further included CBF value, defined as tabular region-averaged CBF measurements extracted from anatomically parcellated perfusion maps. This separation allows the model to independently leverage high-dimensional spatial perfusion patterns and lower-dimensional region-level perfusion metrics.

Beyond neuroimaging, multiple blood-derived modalities were incorporated. Blood metabolites consisted of quantified plasma metabolomic profiles, capturing downstream biochemical activity reflective of systemic metabolic states. Inflammatory markers denote circulating inflammatory biomarker levels measured from plasma, providing insight into immune and inflammatory processes that may influence neurological and systemic health. CBC value included routine clinical laboratory measurements, offering standardized indicators of organ function and physiological status. The Microbiome modality comprised gut microbial taxonomic abundances, representing host–microbe interactions that have increasingly been linked to neuroinflammation and brain function through the gut–brain axis. Finally, the Demographic/clinical data category included demographic and auxiliary structured variables such as age, sex, and additional clinical covariates.

Collectively, this multimodal design enables the model to integrate complementary structural MRI, perfusion-based, metabolic, inflammatory, microbial, and demographic information, thereby capturing both localized neurobiological alterations and systemic physiological states. Such integrative approaches are increasingly recognized as essential for modeling complex brain–body interactions in translational biomedical research.

### H. Model Interpretation: MRI Brain Region Importance Analysis

To investigate the neuroanatomical patterns underlying APOEFormer’s APOE4 predictions, we conducted a brain region importance analysis based exclusively on structural MRI imaging data. This analysis was performed only on MRI-derived features, and did not involve perfusion imaging modalities. The goal was to identify MRI brain regions whose structural perturbation most strongly influences the model’s predictions, while controlling for confounding effects introduced by regional size differences.

For each MRI-derived brain parcel, regional importance was estimated using a masking-based perturbation strategy. Specifically, for a given parcel, a fixed-size subregion was repeatedly masked, and the resulting change in the model’s prediction score was measured. This procedure was repeated 100 times per parcel, and the average change in prediction score was used as the parcel’s importance estimate. By using a constant mask size across all regions, this approach isolates the effect of localized structural perturbations rather than differences in overall parcel volume.

The smallest parcel in the MRI parcellation (“right vessel”) contained very few voxels and produced negligible changes in prediction score, rendering it barely visible in visualization tools such as Mango. Instead, the second-smallest parcel (“right lateral ventricle”) was used to define the reference mask size, ensuring stable and consistent normalization across all MRI regions.

Prior to normalization, white matter regions dominated the importance ranking, as larger and more spatially extensive parcels naturally induced larger prediction changes when masked. This behavior reflects a known limitation of unnormalized perturbation analyses, in which region size strongly biases importance estimates. After applying size-normalized masking, however, the importance distribution shifted toward functionally and clinically relevant cortical regions, revealing a more interpretable pattern of MRI-based contributions.

### I. Model Interpretation: Non-Imaging Feature Importance Analysis

To elucidate the biochemical and physiological signals driving APOEFormer’s APOE4 predictions, we conducted a feature-level importance analysis focusing on non-imaging inputs, including blood-based CBC value profiles and standard blood metabolites measurements. This analysis aimed to identify individual non-imaging features whose perturbation most strongly influences the model’s output and to assess whether the identified signals are consistent with known biological mechanisms implicated in AD.

Feature importance for non-imaging modalities was estimated using a unified masking-based perturbation strategy applied to both metabolomic and blood laboratory data. For each feature, its value was individually masked by setting it to zero while keeping all other input modalities unchanged, and the resulting change in the model’s prediction score for APOE4 status was measured relative to the unmasked baseline. The mean change in prediction score was computed for each feature, yielding a model-agnostic estimate of relative feature contribution.

Perturbation-based feature ablation has been widely used to interpret complex machine-learning models in biomedical applications because it does not rely on internal gradients or assumptions about linearity and remains applicable to heterogeneous architectures [29]–[31]. Applying an identical perturbation protocol across heterogeneous input modalities enables consistent comparison of feature sensitivity within multimodal transformer frameworks.

## III. Experiments And Results

### A. The Accuracy of Predicting APOE4

To evaluate the robustness of APOEFormer under varying data partitions, we conducted 10 independent experimental runs, each using a distinct split of 23 patients in the dataset into training, validation, and test sets. In every run, all three subsets were randomly re-sampled, ensuring that model performance was assessed under diverse data configurations rather than a fixed partition. For each run, the test set consisted of four heldout patients, and predictive performance was measured using classification accuracy for APOE4 carrier status.

The average prediction accuracy on the 10 independent test runs is 75%. Across the 10 tests, APOEFormer achieved test accuracies ranging from 50% to 100% (Figure 2), reflecting variability induced by differences in training, validation, and test compositions. Perfect classification performance (100% accuracy) was observed in four runs (Runs 1, 2, 3, and 5), while 75% accuracy was obtained in two runs (Runs 6 and 7). The remaining four runs yielded accuracies of 50%, corresponding to two correct predictions out of four test samples. Importantly, across the 10 runs, the union of all test sets covered 23 distinct patients, ensuring that evaluation was not restricted to a small subset of individuals. This repeated resampling test strategy allowed each patient to appear in the test set at least once across runs, providing a broader assessment of generalization performance at the subject level rather than reliance on a single held-out cohort.

**Fig. 2.**
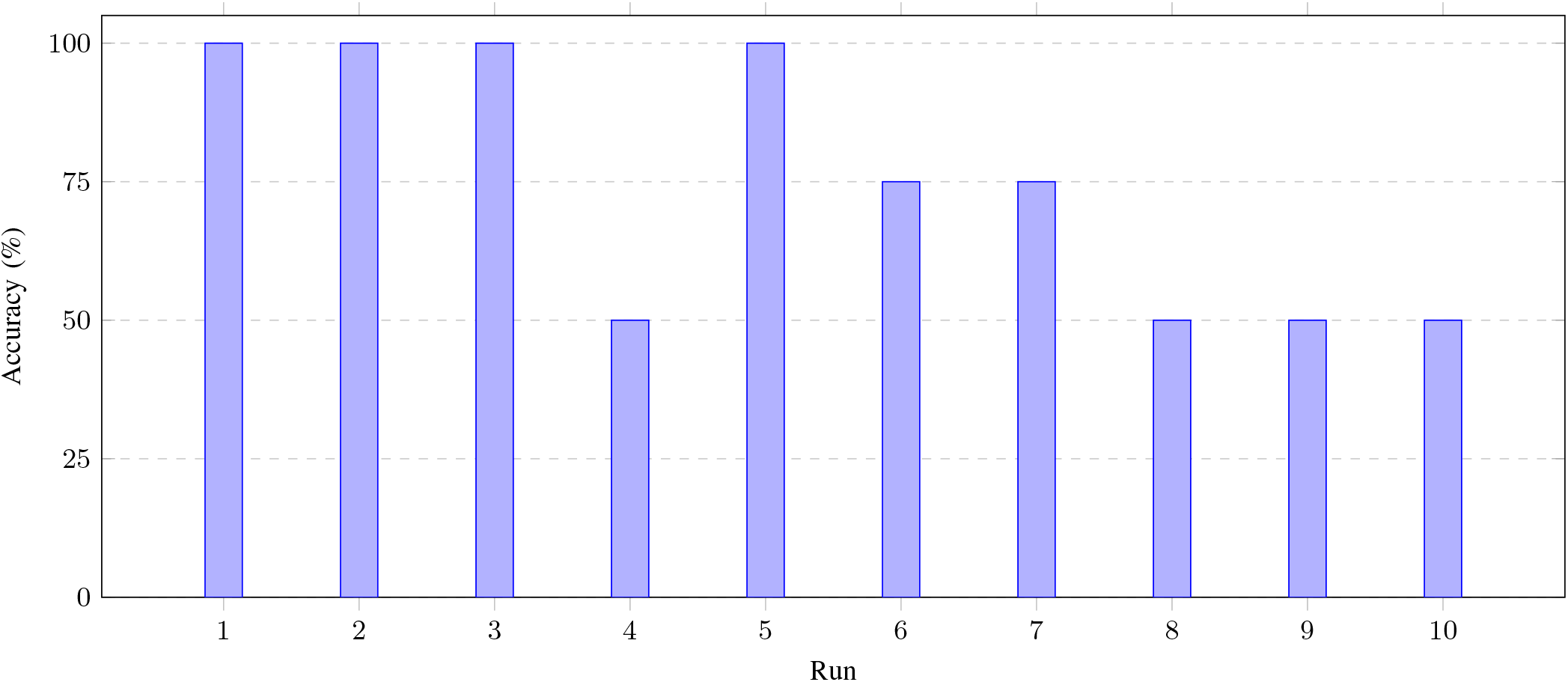
APOE4 classification accuracy across 10 independent test runs. Performance variability reflects differences in data partitioning and sample composition across runs.

Overall, APOEFormer demonstrates good predictive capability for APOE4 carrier-status prediction across multiple independent data splits and across the full cohort of patients, despite limited sample size. The results highlight the importance of repeated evaluation under diverse partitioning schemes in small-sample neuroimaging and genetic prediction studies and motivate further validation on larger cohorts.

### B. Predictive Power of Individual Modalities

To measure the standalone predictive capability of each data type for APOE4 carrier status, we conducted an ablation study to evaluate the predictive performance of APOEFormer using each modality independently. This approach has been widely used in multimodal biomedical learning to disentangle the relative contribution of heterogeneous inputs and to identify dominant sources of predictive signal [8], [10].

The results, summarized in Fig. 3, reveal clear differences in predictive strength across modalities. Blood metabolites achieve the highest standalone accuracy (62%) among all modalities, indicating that circulating metabolic profiles contain strong information related to APOE *ε*4–associated biological processes. This finding is consistent with prior studies demonstrating links between APOE4 genotype and lipid metabolism, mitochondrial function, and systemic metabolic alterations [5], [22].

**Fig. 3.**
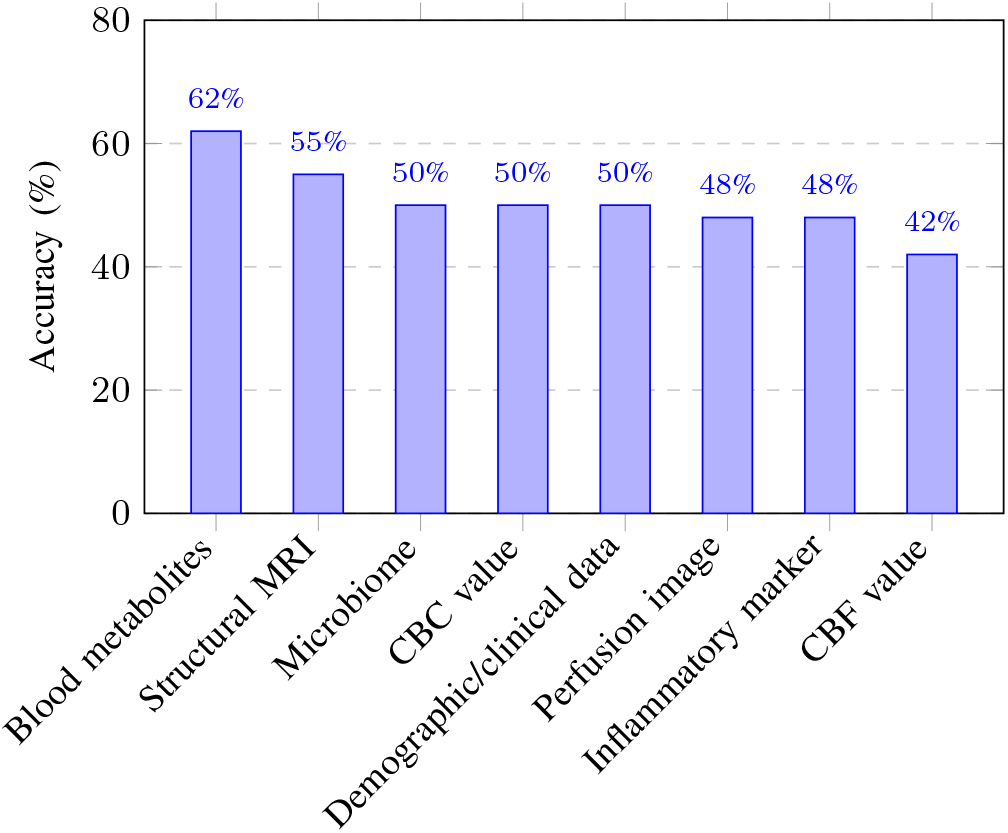
Accuracy obtained when each modality is used individually for model training.

Other blood-based modalities, including CBC value and Inflammatory markers, also exhibit modest predictive performance (accuracy between 48% and 50%). This observation aligns with evidence that APOE4 carriage is associated with altered immune responses and chronic inflammation, which can be reflected in peripheral blood biomarkers [32]. Together, these results highlight the importance of peripheral biochemical and immune-related signals in capturing genetic risk for Alzheimer’s disease.

Structural MRI is the second most predictive feature among all the modalities when evaluated in isolation (accuracy = 55%). Among neuroimaging modalities, it is the most informative, outperforming Perfusion image (accuracy = 48%) and CBF value (accuracy = 42%). Structural MRI has been extensively shown to capture APOE4–related patterns of brain atrophy, particularly in medial temporal and cortical regions, even in cognitively normal individuals [33]. In contrast, Perfusion image and CBF value provide complementary measures of cerebral physiology but exhibit weaker discriminative power when used alone, suggesting that functional alterations may be more subtle or context-dependent.

Microbiome data demonstrates good standalone performance (accuracy = 50%), the same as CBC value. While emerging evidence suggests an association between gut microbiota composition, neuroinflammation, and Alzheimer’s disease risk, these relationships may be influenced by numerous confounding factors [34]. The observed performance is therefore consistent with the microbiome serving as a secondary or modulatory signal rather than a primary driver of APOE4 genetic risk prediction. Demographic/clinical data—including variables such as age, sex, and body mass index (BMI)— demonstrates moderate standalone predictive capability (50%). These variables may capture indirect population-level associations with APOE4 status but lack strong mechanistic specificity when used in isolation. In contrast, CBF value exhibits the lowest accuracy (42%), suggesting limited discriminative power as a standalone modality.

Finally, even though each modality has some predictive power, no single modality fully captures APOE4 status and reaches the 75% prediction accuracy of using all the modalities, supporting the need for multimodal integration.

### C. SHAP-Based Analysis of Modality Importance in Multimodal Integration

To complement the individual modality analysis and gain insight into the model’s decision-making process, we performed a SHAP (SHapley Additive exPlanations) analysis of each modality when they are integrated together. SHAP provides a theoretically grounded framework for attributing model predictions to input features based on cooperative game theory and has been widely adopted for interpreting complex machine-learning models in biomedical applications [35].

As shown summarized in Fig. 4, the SHAP results have some similarity with the trends observed in the ablation study. Blood metabolite data exhibits the highest average SHAP importance (0.449), indicating that it contributes most strongly to the model’s predictions. Other blood-based modalities, including general CBC value (0.357) and inflammatory markers (0.339), also receive relatively high SHAP values, reinforcing their central role in APOE4 classification.

**Fig. 4.**
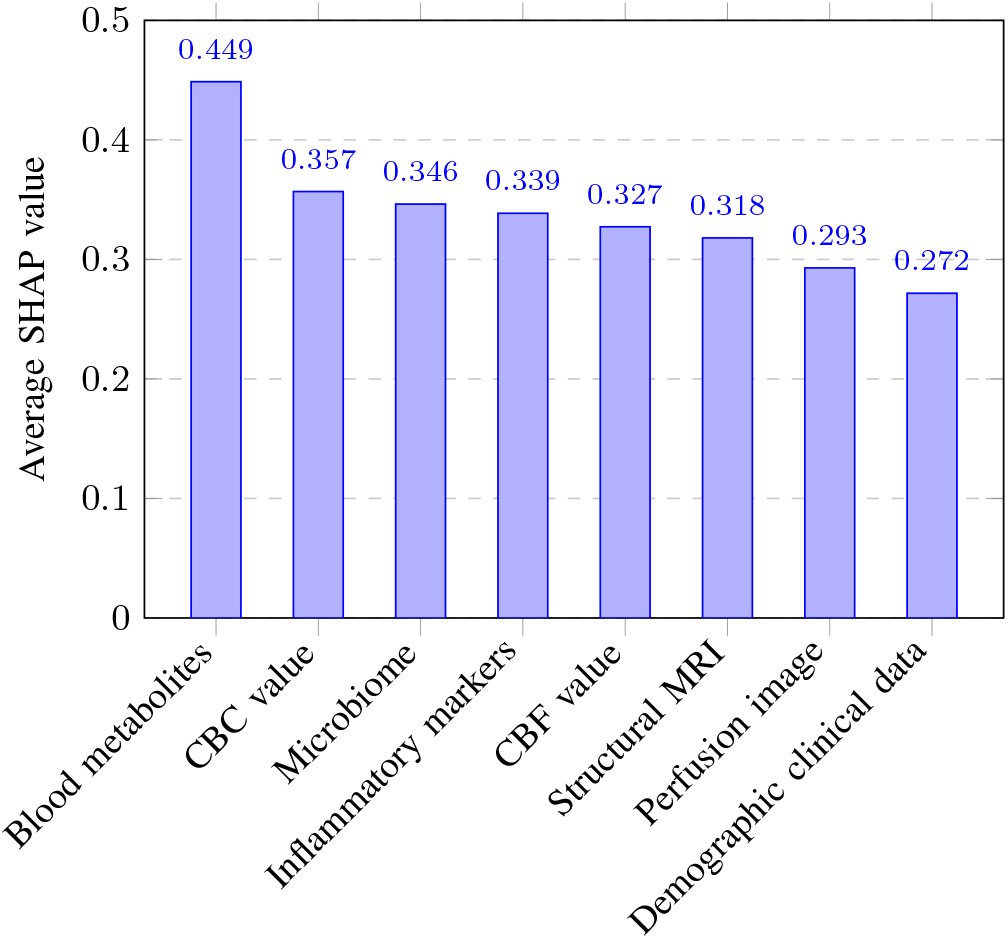
Average absolute SHAP values across input modalities, indicating the relative contribution of each modality to model predictions when they are integrated together.

Microbiome features also show relatively high SHAP importance (0.346), indicating they are complementary with other features in multimodal data integration.

Neuroimaging modalities—including structural MRI, and perfusion imaging—exhibit moderate SHAP importance, lower than that of blood metabolites, CBC value and microbiome. These findings suggest that imaging features play a supplementary role by refining and contextualizing predictions derived from biochemical signals in multimodal data integration, rather than serving as the dominant source of information. This interpretation is consistent with prior multimodal AD studies, where imaging features enhance prediction when combined with molecular or clinical biomarkers [8], [10].

It is worth noting there is some difference in the relative importance of data modalities in the multimodal integration setting (Figure 4) and in the isolation setting (Figure 4). For instance, the relative importance of structural MRI in multimodal data integration is lower than its importance when it is used alone. The difference is because the former consider the interactions between modalities while the latter measure the absolute predictive power of each modality.

Importantly, the results indicate that blood-based modalities—particularly metabolite and inflammatory measurements—constitute the primary source of predictive signal for integrative APOE4 classification, while microbiome features provide complementary context. Structural and functional neuroimaging modalities further enrich the multimodal representation, supporting the rationale for integrating diverse data sources even when individual modalities are only moderately informative.

### D. Identifying Brain Regions Associated with APOE4

Table I summarizes the top-ranked structural MRI brain regions with the highest average absolute importance following feature normalization. Out of nearly 170 anatomical regions evaluated in the structural MRI feature set, only the most influential regions are reported for clarity and interpretability. The top-ranked regions span medial cortical areas, frontal cortex, temporal lobe structures, parietal regions, subcortical nuclei, cerebellar white matter, and interhemispheric white matter tracts. The top five brain regions are also highlighted in Figure 5. The diversity of regions represented among these leading features indicates that model predictions rely on distributed anatomical information rather than localized structural effects.

**TABLE I.** Top brain regions ranked by average absolute importance.

| Region | Average Absolute mean change |
| --- | --- |
| Right Hemisphere White Matter – Isthmus of the Cingulate Gyrus | 0.000769 |
| Right Hemisphere White Matter – Pars Triangularis | 0.000569 |
| Left Putamen | 0.000550 |
| Left Hemisphere White Matter – Temporal Pole | 0.000538 |
| Left Hemisphere White Matter – Rostral Anterior Cingulate Cortex | 0.000494 |
| Left Hemisphere White Matter – Cuneus | 0.000489 |
| Left Hemisphere White Matter – Pars Triangularis | 0.000469 |
| Posterior Corpus Callosum | 0.000440 |
| Left Hemisphere White Matter – Isthmus of the Cingulate Gyrus | 0.000438 |
| Right Hemisphere White Matter – Cuneus | 0.000432 |
| Right Cerebellar White Matter | 0.000414 |
| Right Hemisphere Cortex – Isthmus of the Cingulate Gyrus | 0.000409 |
| Right Hemisphere White Matter – Precuneus | 0.000370 |
| Left Pallidum | 0.000369 |
| Left Hemisphere Cortex – Temporal Pole | 0.000340 |
| Right Thalamus | 0.000339 |
| Right Hemisphere White Matter – Rostral Anterior Cingulate Cortex | 0.000319 |

**Fig. 5.**
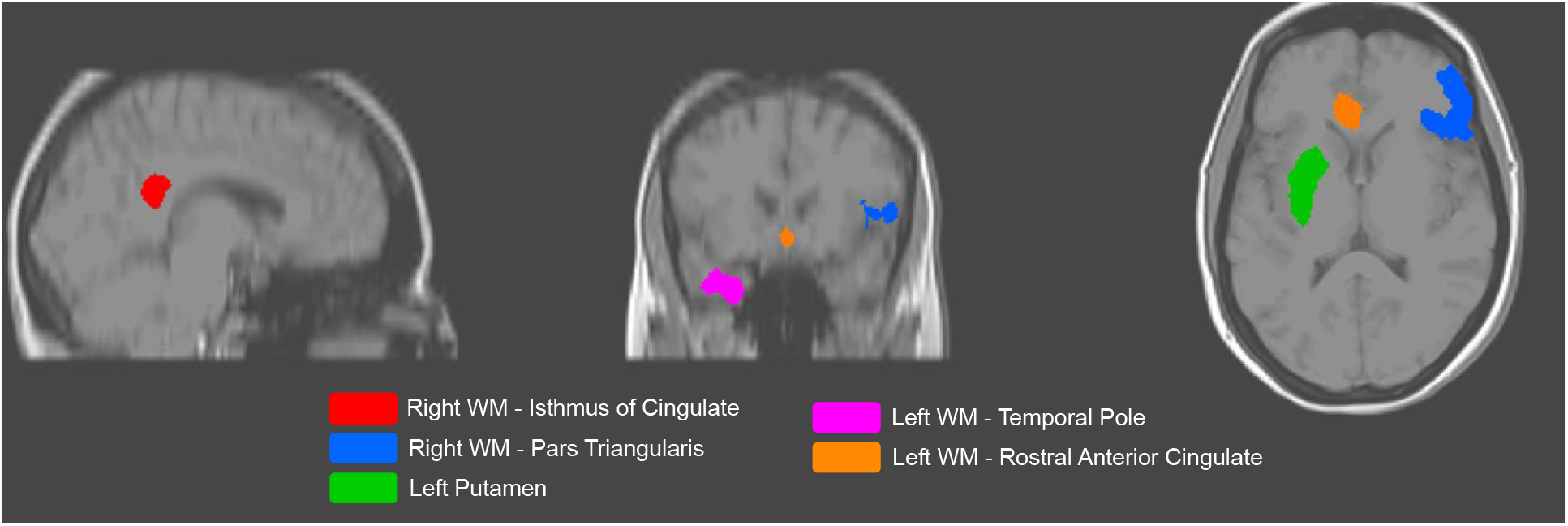
Visualization of the top five structural MRI regions ranked by average absolute mean change (Table I). Highlighted regions correspond to: Right white matter—Isthmus of the cingulate, Right white matter—Pars triangularis, Left putamen, Left white matter—Temporal pole, and Left white matter—Rostral anterior cingulate. Regions are overlaid on a representative participant’s T1-weighted MRI for spatial interpretability.

The isthmus of the cingulate gyrus (wm-rhisthmuscingulate) emerged as the most influential region in the normalized ranking. This region forms part of the medial cortical system and is anatomically positioned at the interface between posterior cingulate and medial temporal areas, supporting integrative cognitive and internally directed processes. Prior neuroimaging work has consistently identified the posterior cingulate and adjacent medial parietal regions as central hubs within large-scale brain networks involved in memory and self-referential cognition [36], [37]. The prominence of the isthmus cingulate in the importance analysis therefore reflects APOE4 prediction sensitivity to structural variation within widely connected medial cortical networks.

Several frontal regions, including the pars triangularis, also ranked among the most influential features. The pars triangularis is a subdivision of the inferior frontal gyrus and has been implicated in higher-order cognitive and language-related functions in functional and structural neuroanatomical studies [38]. Its contribution suggests that frontal cortical morphology may provide complementary predictive information.

Additional regions appearing among the top-ranked parcels include the temporal pole, precuneus, cuneus, thalamus, putamen, pallidum, cerebellar white matter, and posterior corpus callosum. These regions are commonly involved in memory-related processing, visuospatial integration, subcortical–cortical communication, and interhemispheric connectivity in the healthy brain [37]. Their collective presence among influential features further supports the interpretation that the model leverages broad anatomical variation across multiple functional systems.

Across the full MRI parcellation, importance values exhibited a gradual decay rather than a sharp cutoff, indicating that no single region dominates the prediction process. This pattern suggests that APOE4 prediction from structural MRI is driven by coordinated information across multiple brain regions. Feature normalization plays a key role in revealing this distributed structure by mitigating biases related to parcel size and variance magnitude.

Overall, the structural MRI–based importance analysis indicates that model predictions are informed by anatomically meaningful but spatially distributed brain regions. The alignment between regions emphasized by the model and well-established large-scale brain networks supports the interpretability of the MRI component of the framework, while remaining agnostic to specific disease mechanisms or regional causality.

### E. Identifying key blood metabolites for APOE4 prediction

Table II summarizes the top-ranked blood metabolites identified through the perturbation-based importance analysis. The most influential features span diverse biochemical classes, including metabolites related to energy metabolism, amino acid turnover, nucleotide synthesis, and neurotransmission, suggesting that model predictions draw on distributed metabolic information rather than a single dominant pathway.

**TABLE II.** Top metabolites ranked by average absolute importance. Among the 27 quantified metabolites included in the analysis, only the highest-ranked features are shown.

| Metabolite | Mean change |
| --- | --- |
| Creatinine | 0.008487 |
| Inositol | 0.007238 |
| Fructose-6-phosphate | 0.005095 |
| Threonine | 0.004370 |
| Succinate | 0.003907 |
| Uridine | 0.003879 |
| Proline | 0.003756 |
| Pterin | 0.003738 |
| Erythrose | 0.003425 |
| Glycine | 0.003168 |
| Gamma-aminobutyrate | 0.003141 |
| Tyrosine | 0.003030 |
| Glycerol | 0.002733 |
| Tryptophan | 0.002499 |
| Methionine | 0.002415 |
| Malate | 0.002377 |

Creatinine exhibited the largest average perturbation effect. As a downstream product of creatine metabolism, circulating creatinine is commonly interpreted as a marker of systemic energy metabolism and renal clearance rather than a direct indicator of neural function. Prior studies have reported associations between altered brain energy metabolism, mitochondrial efficiency, and neurodegenerative processes, including AD, particularly in populations at elevated genetic risk [39]. In this context, the influence of creatinine in the model likely reflects broader metabolic or physiological states correlated with disease risk, rather than a specific mechanistic role.

Inositol also ranked among the most influential metabolites. Inositol participates in phosphatidylinositol signaling pathways that regulate membrane dynamics and intracellular signaling across many tissues. Altered inositol levels have been observed in neuroimaging and biochemical studies of neurodegenerative conditions, often interpreted as reflecting glial activity or changes in cellular signaling environments. However, such alterations are not specific to AD and should be viewed as indicators of general metabolic or cellular state rather than disease-specific markers.

Fructose-6-phosphate, a central intermediate in glycolysis and related biosynthetic pathways, showed a notable contribution to model predictions. Disruptions in glucose metabolism and reduced cerebral glucose utilization are well-documented features of AD at the systems level, including in asymptomatic individuals at genetic risk [39]. The importance of glycolytic intermediates in the model is therefore consistent with prior observations of metabolic vulnerability in neurodegeneration, while remaining nonspecific to brain tissue or pathology.

Additional metabolites with elevated importance—including succinate, malate, glycerol, and erythrose—are involved in mitochondrial respiration and intermediary metabolism, while amino acids such as glycine, threonine, proline, methionine, tyrosine, and tryptophan reflect protein turnover and neurotransmitter precursor availability. Nucleotide-related metabolites such as uridine further reflect cellular maintenance and biosynthetic activity. Broad alterations in mitochondrial metabolism, amino acid homeostasis, and neurotransmitter balance have been reported in AD and related neurodegenerative disorders, although these changes are typically systemic and multifactorial rather than disease-specific [40], [41].

### F. Identifying key CBC value measurements for APOE4 prediction

Table III reports the most influential blood-based laboratory variables as quantified by the perturbation-based mean change analysis. The identified features span renal, hepatic, metabolic, hematological, and immune-related processes, underscoring the contribution of systemic physiological states to APOE4 prediction and AD–related risk modeling.

**TABLE III.**
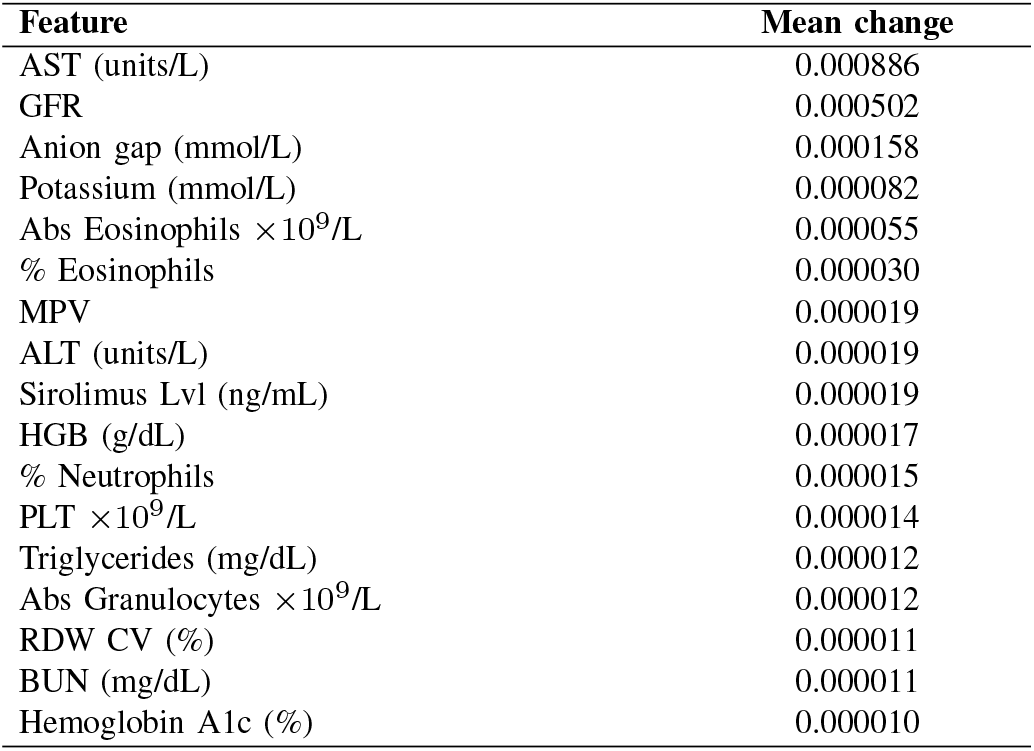
Top blood laboratory features ranked by perturbation-based mean change. Among the 46 laboratory variables included in the analysis, only the highest-rankedfeatures are shown.

Markers of renal and metabolic function, including estimated glomerular filtration rate (GFR), blood urea nitrogen (BUN), and the anion gap, ranked among the most influential variables. Impaired renal function and metabolic imbalance are increasingly recognized as contributors to cognitive decline, potentially through shared vascular pathology, chronic inflammation, and toxin clearance dysfunction. Epidemiological and clinical studies have reported associations between reduced kidney function and increased risk of dementia and AD, supporting the relevance of these systemic markers in neurodegenerative risk stratification [42].

Liver-associated enzymes, particularly aspartate aminotransferase (AST) and alanine aminotransferase (ALT), also exhibited measurable influence. Although these enzymes are not specific to central nervous system pathology, they serve as indicators of systemic metabolic status and mitochondrial stress. Growing evidence suggests that metabolic dysregulation and mitochondrial dysfunction play important roles in AD pathophysiology, including impaired energy metabolism, oxidative stress, and neuronal vulnerability [40], [43].

Several hematological indices—including hemoglobin concentration, platelet count (PLT), red cell distribution width (RDW), and mean platelet volume (MPV)—were likewise influential. Alterations in blood cell morphology and platelet activity have been linked to cerebrovascular dysfunction and systemic inflammation, both of which are implicated in AD progression. In particular, elevated RDW and abnormal platelet activation have been associated with cognitive impairment and neurodegenerative disease in prior studies [44].

Immune-related measures, including eosinophil, neutrophil, and granulocyte counts, further highlight the role of peripheral immune activity. Chronic systemic inflammation is a well-established contributor to AD, influencing microglial activation, amyloid pathology, and neurodegenerative cascades. Importantly, inflammatory processes interact with APOE4–associated risk mechanisms, amplifying susceptibility to neurodegeneration [45], [46].

These results demonstrate that APOEFormer captures distributed signals across multiple physiological systems rather than relying on isolated biomarkers. The integration of standard clinical laboratory features with metabolomic and imaging-derived representations aligns with the multifactorial and systemic nature of AD, supporting the use of multimodal learning frameworks for robust modeling of genetic phenotypes.

## IV. Discussion

In this study, we introduce APOEFormer, a multimodal transformer for predicting APOE4 carrier status through the integration of heterogeneous biomedical data, including structural brain MRI, Perfusion image, CBF value, blood-based biomarkers, gut microbiome profiles, and auxiliary demographic/clinical variables. APOEFormer adopts a two-stage learning paradigm in which modality-specific encoders are first pretrained and aligned via contrastive representation learning, followed by multimodal integration using a transformer architecture for subject-level prediction. Transformer-based attention enables flexible modeling of cross-modal dependencies without imposing a fixed ordering over modalities, making it particularly well suited for heterogeneous biomedical inputs where modality order is not inherently defined [16], [47].

Across 10 independent experimental runs, APOEFormer achieved a mean classification accuracy of 75% for APOE4 carrier prediction on the test data. Importantly, the union of test sets across runs covered all 23 subjects in the cohort, ensuring comprehensive subject-level evaluation rather than reliance on a single held-out subset. This repeated evaluation protocol mitigates variance arising from small sample sizes and has been recommended as best practice in neuroimaging studies with limited cohorts [48], [49]. Although performance varied across individual runs due to phenotypic heterogeneity and small test sizes, the consistency of results across diverse data splits suggests that the transformer does not overfit to a particular partition and is capable of generalizing across individuals.

A central finding of this work is the importance of multimodal biological integration for APOE4 prediction and characterization. Both modality-wise ablation experiments and SHAP-based interpretability analyses consistently identified blood-based modalities—particularly metabolite markers—as the strongest contributors to predictive performance. This observation aligns with prior evidence linking APOE4 to systemic metabolic dysregulation, altered lipid metabolism, immune activation, and chronic inflammation [22], [50]. Structural MRI emerged as the second most informative imaging modality when evaluated independently. Rather than acting as the primary driver of predictions, imaging features provided complementary anatomical context that refined predictions derived from biochemical signals. Microbiome and auxiliary clinical variables exhibited also contributed meaningfully to the prediction when used in isolation or within the transformer-based multimodal fusion setting.

Perturbation-based analyses revealed that influential features span energy metabolism, amino acid turnover, mitochondrial intermediates, renal and hepatic function, hematological indices, and immune-related markers, indicating that predictive signals arise from distributed systemic physiology rather than isolated biomarkers.

At the metabolite level, highly ranked features included creatinine, inositol, glycolytic intermediates, tricarboxylic acid cycle metabolites, amino acids, and neurotransmitter-related compounds. These results suggest that global metabolic state—including energy utilization, mitochondrial function, and biosynthetic activity—provides meaningful context for genetic risk modeling. Importantly, the model does not rely on any single metabolite, but instead integrates coordinated signals across multiple metabolic pathways, consistent with the multifactorial and systemic nature of neurodegenerative vulnerability.

Complementary patterns were observed in blood-based laboratory features, particularly within the CBC value modality, where markers of renal function, hepatic metabolism, hematological status, and immune activity contributed to predictive performance. The prominence of measures such as glomerular filtration rate, liver enzymes, red blood cell indices, platelet-related metrics, and inflammatory cell counts further supports the interpretation that peripheral physiological states reflect biologically relevant context for APOE4–associated risk. These findings align with prior evidence that metabolic dysregulation, vascular health, and immune activation interact with genetic susceptibility in AD, while remaining nonspecific to any single disease mechanism.

Despite the encouraging performance and interpretability results, several limitations of the proposed framework should be acknowledged. First, the relatively small cohort size restricts statistical power and increases variance in performance estimates, even when using repeated data splits. Although the repeated evaluation protocol mitigates overfitting to individual partitions, performance metrics should be interpreted cautiously and validated in larger, independent cohorts.

Second, the perturbation-based interpretability analyses quantify feature influence within the trained model but do not establish causal relationships between specific biological variables and APOE4 carrier status. Highly ranked metabolites and blood laboratory measures should therefore be interpreted as correlates rather than mechanistic drivers, and further experimental or longitudinal studies are required to disentangle causality.

## Data Availability

The data used in this study are not publicly available due to privacy and ethical restrictions but are available from the corresponding author upon reasonable request and with appropriate approvals.

